# Estimates of the impact on COVID-19 deaths of unequal global allocations of vaccines

**DOI:** 10.1101/2022.01.26.22269347

**Authors:** J. Paul Callan

## Abstract

During 2021, COVID-19 vaccinations were delivered much more rapidly in some countries than in others. Ethical principles would have suggested allocating available vaccines to people by age, irrespective of where they live, because mortality risks from COVID-19 are much higher for older people. The World Health Organization recommended initial allocations of vaccines to countries based on their total population size, in part due to uncertainty about how COVID-19 would affect different countries.

This paper estimates how many people would have died from COVID-19 up to 31 October 2021 if either of these allocation rules had been applied, compared to estimates of actual COVID-19 deaths. The estimates suggest that allocating vaccines by age would have resulted in between 500,000 and 1,500,000 fewer deaths globally (with a best estimate of 1,090,000 fewer deaths), while allocating vaccines between countries based on national population sizes would have reduced total deaths globally by between 450,000 and 2,100,000 (with a best estimate of 1,440,000 fewer deaths).

Most low-and middle-income countries would have seen reductions in deaths, with the greatest absolute numbers in large middle-income countries (especially Bangladesh, India and Indonesia). More deaths would have taken place in many high-income countries, with the greatest absolute numbers in the United States and Turkey, and the greatest percentage changes in Arabian Peninsula countries, Israel and some island states. In most European Union countries, deaths would not have differed much if vaccines were allocated by age, because they would have received more vaccine doses during the early months of 2021 but fewer later in the year.

Although allocation of vaccines by age should intuitively lead to fewest deaths, the estimated deaths would have been even lower if vaccines were allocated based on population size. Allocation by population would have directed disproportionate numbers of vaccines to a set of countries – especially India, Bangladesh and Indonesia – which experienced large outbreaks due to the Delta variant in 2021 after having previously limited infections through “ flattening the curve”.

Sequencing of vaccination by age in national vaccination rollouts is critical to maximizing the numbers of lives saved. The estimated gains from fairer global vaccination allocation would be greater if high-income and upper-middle-income countries did not sequence vaccinations by age cohort, and would be lower if lower-middle-income and low-income countries did not vaccinate most of their elderly before the general population, whether due to policy choices or people not accepting vaccines made available to them or logistical difficulties in vaccine delivery.

The estimates correspond to a reduction of between 8.5% and 10.7% (with a best estimate of 10.4%) of the total estimated actual deaths from COVID-19 between 1 January and 31 October 2021, if vaccines were allocated by age, and between 7.8% and 15.0% (with a best estimate of 13.6%) of total estimated actual deaths, if vaccines were allocated based on national population sizes. These percentages are small, despite the large differences in vaccine deployment between countries, because the mostly high-income countries which vaccinated their populations faster have disproportionately large numbers of elderly people. If a future SARS-CoV-2 variant, or a future pandemic, were to have fatality rates that are similar across age groups, or that are higher for children and young adults, then unequal global allocation of vaccines would have a much more severe effect on overall global mortality than it has for COVID-19 so far.

## Introduction: Alternative approaches to allocation of vaccines among countries

COVID-19 vaccination proceeded rapidly after the first vaccines received emergency use authorizations in December 2020. According to the Our World In Data COVID-19 vaccination dataset *[1]*, up to 31 December 2021, 9.17 billion vaccine doses were administered, amounting to 116.46 doses per 100 people worldwide. More than 58.0% of the world’s population had received at least one dose of a vaccine, and 49.4% of people were fully vaccinated. However, rollouts of vaccines were extremely uneven. As shown in Figure 1, vaccinations proceeded at radically different paces in different countries. Most Africans remained unvaccinated, with only 22.50 doses delivered per 100 people up to the end of 2021.

**Figure 1.**
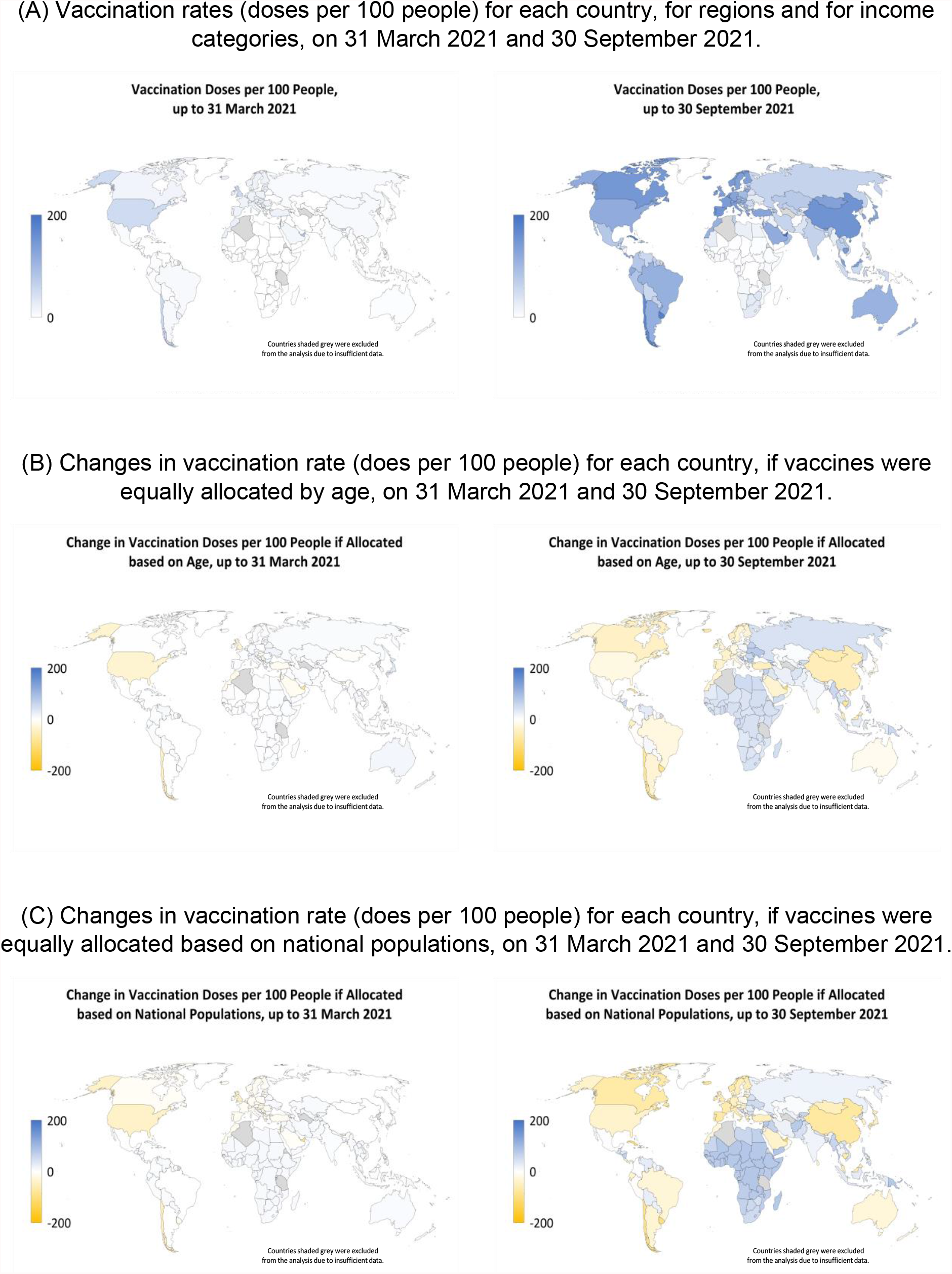
Actual COVID-19 vaccination rates and deviations from more ethical allocation scenarios.

The allocation of vaccines among countries has deviated from ethical principles. According to medical ethicists, including Emanuel et al. *[2]*, “ maximizing saving lives” is most important in a pandemic, and thus “ justifies giving older persons priority for vaccines immediately after health care workers and first responders,” because “ Covid-19 outcomes have been significantly worse in older persons.” Following these ethical principles, policies within most countries *[3]*, and guidelines from the World Health Organization *[4]*, call for deployment of available vaccines first to healthcare workers and then to old and vulnerable people, starting with the oldest groups. Across countries, these principles suggest allocating vaccine doses to countries in such a way as to allow different countries to vaccinate their healthcare workers, and then successive age cohorts, starting with the eldest, at the same time as in all other countries – although Emanuel et al. *[5]* point out that “ it is an empirical question whether this prioritization optimally reduces death”.

An alternative rule for allocating vaccines across countries is to share doses based solely on each country’s population size, not taking age into account. This rule was used by COVAX, the global vaccines facility (led by CEPI, Gavi and WHO) through which many developing countries receive most of their vaccines, for its initial deliveries, and was also used by the European Union in allocating vaccine doses among its member states. The allocation-by-population rule was justified for the COVAX facility for reasons which include (1) “ uncertainty about … the course of the pandemic in different regions” and (2) a desire to “ provide certainty to all countries … that they will receive a sizeable number of vaccine doses” *[6]*. Another argument in favour of this rule is that mortality risks for people of all ages are likely higher in low-and middle-income countries, due to poorer healthcare, which could compensate partly for the younger populations in those countries. However, Emanuel et al. *[5]* criticize this allocation rule because “ a fair distribution of COVID-19 vaccines should respond to the pandemic’s differential severity in different countries”.

Figure 1 illustrates how much different the vaccination rates would have been in each country, region and income category, if each of these two rules for allocating vaccines across countries – by age and based on national population size – had been applied, instead off the actual allocations which were driven largely by “ vaccine nationalism”.

## Question: How many lives, if any, would fairer vaccine allocation have saved

What have been the consequences of the unethical distribution of vaccines among countries? Specifically, how many fewer (or more) people would have died from COVID-19 if vaccines were distributed more fairly?

## Methods: Approach used to estimate the effects of alternative vaccination scenarios

### Simple calculations provide estimates to answer these questions

In these calculations, we estimate, for each of 183 countries and territories *[7]* (referred to as “ countries”), home to 98% of the world’s population, and for each day of the pandemic, the overall average infection fatality ratio (IFR), i.e., the number of deaths expected for every infection, given the actual vaccination rates. Next, we estimate what the overall average IFR would have been, for each country and each day, under each of the two alternative scenarios for allocation of vaccines globally among countries – namely (1) allocation by age and (2) allocation based on national populations.

From the ratio between the overall average IFR in the alternative scenarios and the estimated actual overall IFR, we can estimate what the reported deaths from COVID-19 *[8]* would have been under the alternative scenarios. Finally, we estimate real numbers of deaths – in the actual and alternative vaccine allocation scenarios – from the reported numbers of COVID-19 deaths, using estimates of the undercounting of COVID-19 deaths in each country (largely drawn from excess mortality estimates by The Economist *[9]*).

The calculations of overall average IFR estimates utilize several other parameters. They use numbers published by the United Nations for populations by age cohort in each country *[10]*. They use published estimates for IFRs for people of different ages *[11]* (assumed to apply across countries), and for vaccine effectiveness in reducing mortality *[12]*. Based on the announced policies of various countries *[3]*, they assume that vaccinations in high-income countries (HICs) and upper-middle-income countries (UMICs) are sequenced by 5-year-wide age cohorts, starting with the oldest, up to an assumed maximum uptake in each cohort, and that vaccinations in lower-middle-income countries (LMICs) and low-income countries (LICs) are provided first to people above a given threshold age, and then to people below the threshold age, up to an assumed maximum uptake for the groups above and below the threshold age. Sensitivity analyses were conducted to test how the results changed upon varying these parameters and assumptions.

The calculations make estimates for deaths up to 31 October 2021, which are influenced by vaccinations administered up to around the end of September 2021 because vaccines take 7 to 14 days to generate full immunity to the virus *[13]* and deaths occur at an average of 20-25 days after infection *[14]*. There are two reasons to focus on vaccinations up the end of September 2021 and deaths up to the end of October 2021. First, for most countries, the total numbers of vaccine doses administered were determined by the availability of vaccines up to that date, but the demand for vaccination became the determining factor for total vaccine doses in increasing numbers of countries after that date. Second, booster shots were relatively rare prior to 30 September 2021 – accounting for only 0.47 doses per 100 people up to that date – but have become more common since then.

The Model Description, annexed to this article, provides a full specification of the calculations and sensitivity analyses.

## Findings: Allocating vaccines by age would have saved an estimated 1,090,000 people, while allocating by population would have saved an estimated 1,440,000 people

If vaccines had been allocated based on age, we estimate that 1,090,000 fewer deaths from COVID-19 would have occurred globally up to the end of October 2021 – 14.96 million deaths compared to the estimated 16.06 million deaths which actually occurred up to that date. Thus, the death toll from COVID-19 would have been 6.8% lower over the entire pandemic up to 31 October 2021, or 10.4% lower for the period between 1 January and 31 October 2021 (i.e., roughly the time during which vaccines were available). Figure 2 shows how deaths would have decreased or increased, for various countries and groups of countries, under the allocation-by-age scenario compared to the actual allocation of vaccines. The figure shows these differences as absolute numbers of deaths and as a percentage of estimated actual deaths from COVID-19 between 1 January and 31 October 2021. In all, 139 countries would have had reduced numbers of deaths, under the allocation-by-age scenario, while 40 countries would have seen more deaths. (Four of the 183 countries and territories considered, together with many of those not included in these calculations, are small island states and territories in which there have been no or few reported deaths from COVID-19.)

**Figure 2.**
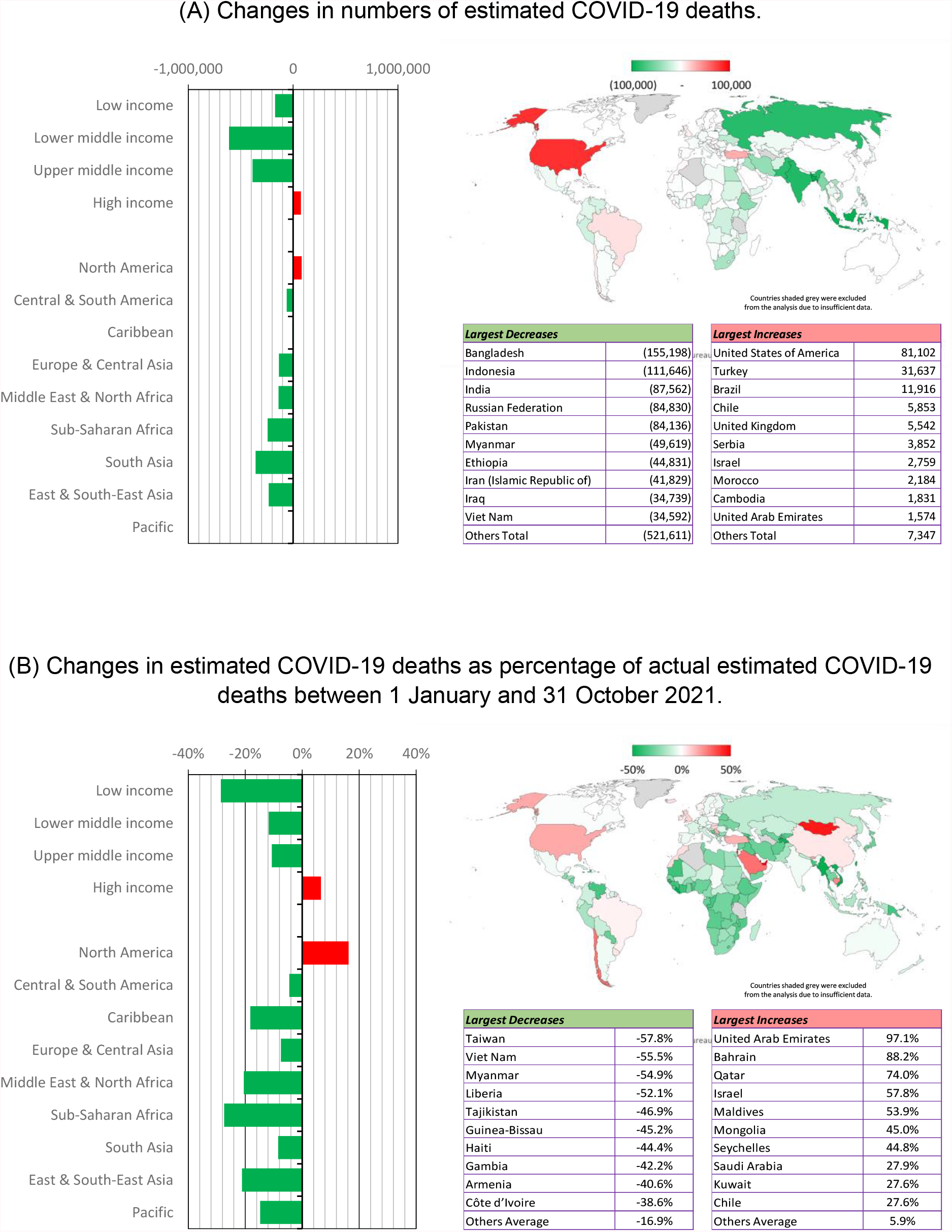
Changes in estimated COVID-19 deaths if vaccines were equally allocated based on age, compared to estimated actual COVID-19 deaths, up to 31 October 2021.

If vaccines had been allocated to countries in proportion to their total populations, not taking age into account, we estimate that 1,440,000 fewer deaths from COVID-19 would have occurred globally up to the end of October 2021 – 14.62 million deaths compared to the estimated 16.06 million deaths which actually occurred up to that date. The death toll from COVID-19 would have been 9.0% lower over the entire pandemic up to that time, or 13.6% lower for the period between 1 January and 31 October 2021. Figure 3 shows how deaths would have decreased or increased, for various countries and groups of countries, under the allocation-by-population scenario compared to the actual allocation of vaccines. In this scenario, 116 countries would have had reduced numbers of deaths, while 64 countries would have seen more deaths.

**Figure 3.**
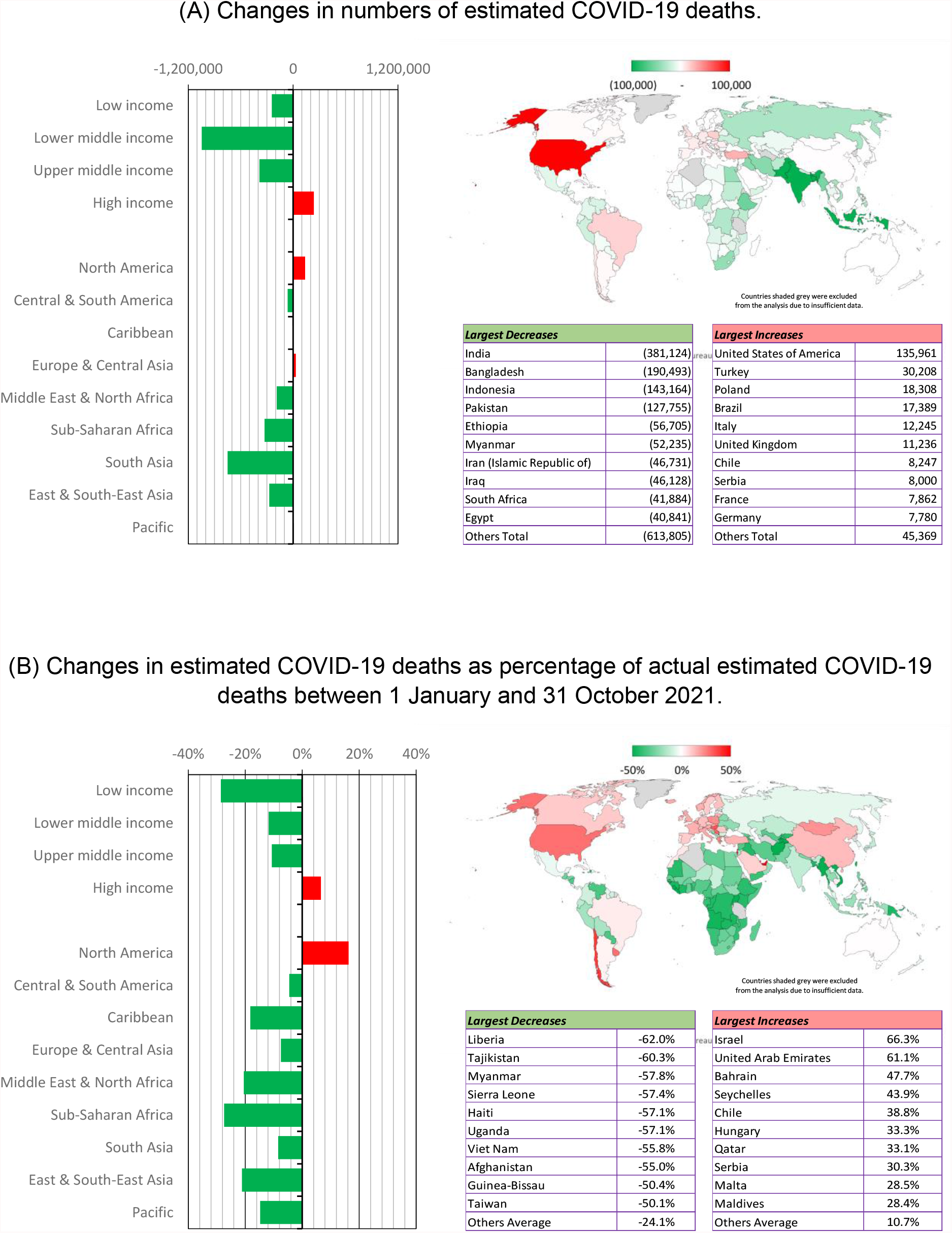
Changes in estimated COVID-19 deaths if vaccines were equally allocated based on total national populations, compared to estimated actual COVID-19 deaths, up to 31 October 2021.

Most of the lives saved if vaccines had been allocated more fairly – either by age or based on total populations – would have been in large middle-income countries, with the largest numbers in Bangladesh, India and Indonesia. The percentage reductions in deaths from COVID-19 would have been greatest, however, in low-income countries, most in Africa, where actual vaccination rates were the lowest, as well as in Taiwan, Viet Nam and Myanmar, where vaccinations only ramped up significantly in mid-2021 after Delta outbreaks had already started. As a share of their populations, the lives saved in low-income and lower-middle-income countries would have been roughly similar; low-income countries had more drastic gaps in vaccination compared to lower-middle-income countries, but low-income countries also have the youngest populations.

Some countries would have seen increased deaths under the fairer-allocation scenarios – 40 out of 183 countries if allocating by age and 65 out of 183 countries if allocating based on total populations. However, the number of extra lives lost in these countries would have been much fewer than the number of extra lives saved in the rest of the world. High-income countries, together with some upper-middle-income countries, account for most of the countries in which deaths would have increased. The greatest increases in absolute numbers would have been in the United States and Turkey, while the greatest percentage increases in COVID-19 deaths would have been in Arabian Peninsula countries, Israel and some island states.

It is perhaps surprising that estimated deaths would have been lower if vaccines were allocated based on population size rather than based on age, considering how steeply mortality rates increase with age *[11]*. The reason is that allocation-by-population would have directed disproportionate numbers of vaccines to a set of countries – most of all India, Bangladesh, and Indonesia – which experienced large outbreaks due to the Delta variant in 2021 after having previously limited infections through strongly “ flattening the curve” *[15]*.

Most European Union countries would have experienced small reductions in numbers of deaths if vaccines had been allocated by age; most would have experienced extra deaths if vaccines had been allocated to countries based on total population size. If vaccines had been allocated by age, most of Europe would have had more vaccines in the early months of 2021, reducing mortality during Alpha-variant-driven waves, but would have had fewer vaccines later in the year, increasing mortality during the Delta-driven waves.

Countries which have kept COVID-19 infections to very low levels throughout 2021 – including Australia, Bhutan, China, Iceland, Japan, Mongolia, New Zealand, Norway, South Korea, and many small island states – show little difference in numbers of deaths across the scenarios, because the timing of vaccinations does not greatly affect the estimated number of deaths. On the other hand, for countries in South-East Asia, which largely kept COVID-19 out until the emergence of the Delta variant, the timing of vaccination was critical. Cambodia, which vaccinated people faster than the world average, has experienced few deaths, and would have experienced more under the fairer allocation scenarios. Thailand and Viet Nam would have saved lives had they been able to vaccinate more people, especially the elderly, prior to their outbreaks of the Delta variant in mid-2021.

## Sensitivity Analyses: Fairer global allocation of vaccines would have saved lives for all plausible parameters; sequencing vaccinations by age within countries is critically important to maximize lives saved

Although the scenarios and calculations are highly simplified, substantial improvements in COVID-19 mortality are found under the allocation-by-age and allocation-by-population scenarios under a wide range of possible assumptions for the parameters used in the calculations. Figure 4 presents the range of values for differences in COVID-19 mortality calculated in a series of sensitivity tests, which altered the assumptions for each of the major parameters. Details of the sensitivity analyses are provided as part of the Model Description.

**Figure 4.**
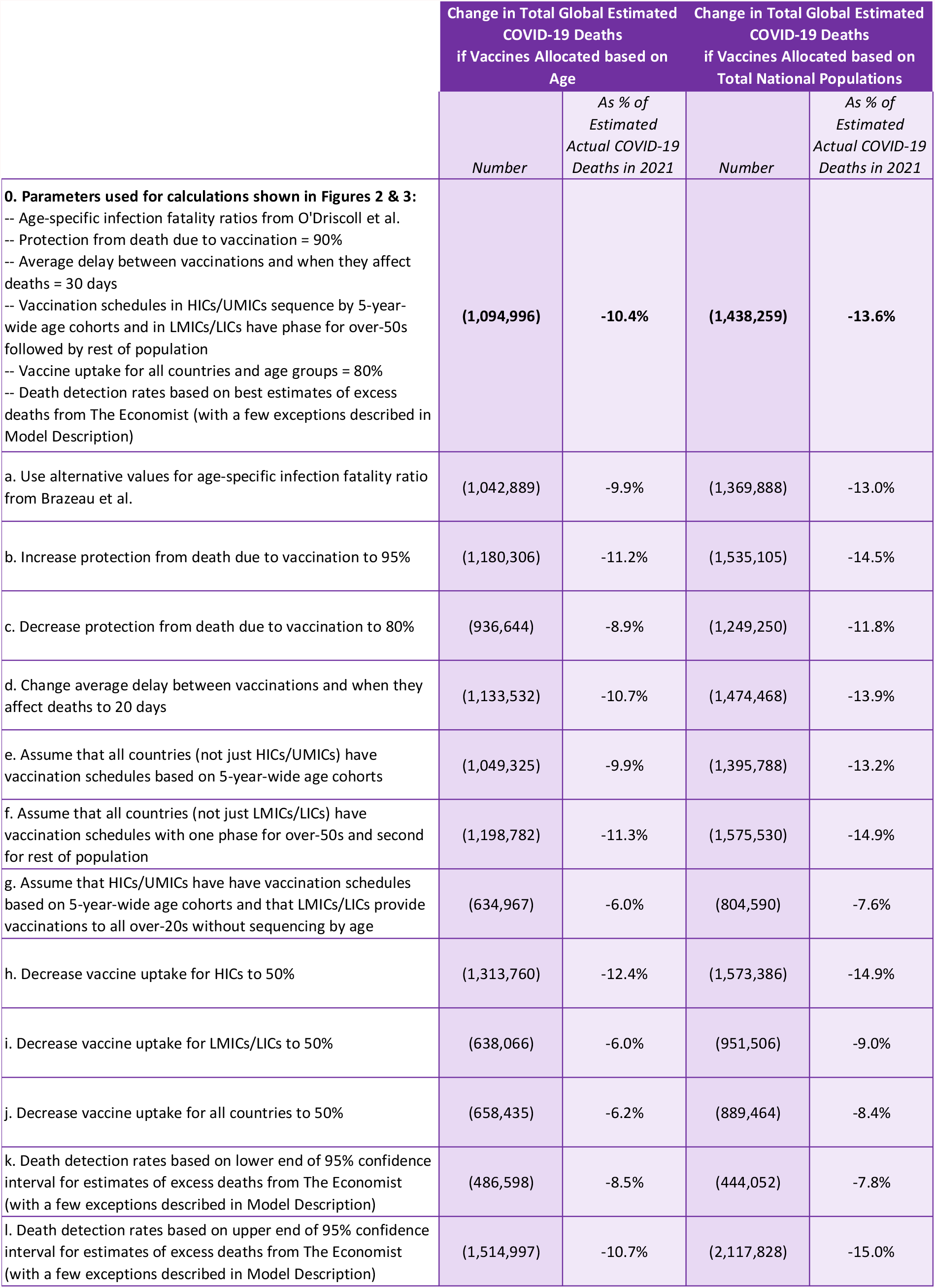
Changes in estimated COVID-19 deaths under the alternative global vaccine allocation rules, for different assumptions regarding key parameters in the calculations.

The sensitivity analyses illustrate the vital importance of sequencing vaccinations by age and of ensuring high uptake to make the optimal use of available vaccines.

Estimates for extra lives saved under alternative scenarios would be lower if vaccination schedules in LMICs and LICs are assumed to be less strongly sequenced by age (e.g., having a threshold age for the first phase of vaccinations that is lower than the 50 years assumed for the calculations whose results are shown in Figure 2 and 3, or moving to vaccination of the general population before vaccinating all of the older and vulnerable people who wish to be vaccinated). Conversely, the estimates for extra lives saved under the alternative scenarios, as a percentage of actual COVID-19 deaths, would be greater if vaccination schedules in HICs and UMICs are assumed to be less strongly sequenced by age (e.g., having one phase for all elderly people instead of sequencing by 5-year-wide age cohorts as assumed for the calculations whose results are shown in Figures 2 and 3). In practice, there were many UMICs and some HICs that did not sequence vaccinations by 5-year-wide or 10-year-wide age cohorts *[3]*, but there is limited information on actual vaccination schedules in most LMICs and LICs.

The numbers of lives saved under the alternative scenarios would be reduced if vaccination uptake is lower (especially among older people) in LMICs/LICs than in HICs/UMICs, and higher if vaccination uptake is lower (especially among older people) in HICs. The vaccination uptake parameter in these calculations could be reduced due to people not accepting vaccines made available to them or due to logistical difficulties in vaccine delivery, either of which could cause countries to offer vaccines to lower age groups before completing vaccinations for older age groups. Countries may differ in rates of vaccine refusal – for example vaccination rates are low in many countries Eastern Europe despite availability *[1]* – but experience to date and surveys on vaccine hesitancy do not suggest systematic differences between countries in different income categories *[16]*. It is possible that logistical difficulties may lead to delays in vaccinating priority groups, and to vaccination programs moving to lower age groups before completing older groups, in some LMICs and LICs, but information is limited at the present time.

Note that the calculations assume that changes in global allocation of vaccine doses among countries would not have led to changes in the sequencing of vaccination programs within individual countries.

The calculations presented here consider only the effect of vaccination in reducing mortality from COVID-19 – including both people who avoid infection completely if “ attacked” by the SARS-CoV-2 virus and people who experience milder disease than if they were not vaccinated. The calculations assume that the dynamics of disease transmission would not have changed much in the different scenarios, and specifically that similar numbers of people would have been “ attacked” by the virus. This assumption is reasonable for two reasons. First, while vaccination appears to have curtailed virus transmission to some degree, it does not seem to have substantially altered the disease dynamics in many countries, especially after the high-transmissibility Delta variant started to circulate *[17]*. Second, it is likely that many countries might have strengthened or relaxed their social distancing measures if disease transmission had been faster or slower due to higher or lower vaccination rates than those which actually occurred. If vaccines could have altered disease dynamics, sharing vaccines equally by age or by population would likely have resulted in even fewer deaths in those countries that would have received more vaccines earlier, and might have increased deaths in countries that would have received fewer vaccines – but both effects would likely be small in comparison to the estimated changes in deaths presented here.

## Concluding Remarks: Inequalities in allocation of vaccines between countries resulted in greater numbers of deaths from COVID-19 – and would have cost even more lives if COVID-19 affected the young as much as the old

Fewer people would have died from COVID-19, worldwide, if the world had made more ethical choices in allocating vaccine doses across countries. Allocating vaccines based on age would have saved between 500,000 and 1,500,000 more lives, with a best estimate of 1,090,000 fewer deaths, while allocating vaccines based on total national populations would have saved between 450,000 and 2,100,000 more lives, with a best estimate of 1,440,000 fewer deaths.

These estimates represent between 8.5% and 10.7% (best estimate 10.4%), and between 7.8% and 15.0% (best estimate 13.6%), respectively, of the total estimated deaths from COVID-19 between 1 January and 31 October 2021.

It is perhaps surprising that vaccine nationalism did not cause even more deaths compared to more ethical vaccine allocation rules, considering the drastic gaps in vaccine deployment between countries. The reason is that countries which vaccinated more people, and did so earlier, are mostly high-income countries which have disproportionately large numbers of elderly people who are most vulnerable to COVID-19. If future SARS-CoV-2 variants, or a future pandemic, have fatality rates that are similar across age groups, or that are higher for children and young adults, then unequal allocation of vaccines would have a much more severe effect on overall global mortality than it has had for COVID-19 so far.

## Annex: Model Description

We consider three scenarios for the global allocation of vaccines across countries: *Actual Allocation (AA)*, which is essentially a “ vaccine nationalism” scenario, *Equal Allocation by Age (EAA)*, and *Equal Allocation by Population (EAP)*.

In any scenario for allocation of vaccine doses across countries and for how vaccinations are sequenced within a country, on any given day, the overall infection fatality ratio (IFR) for a national population can be determined through the following formula:

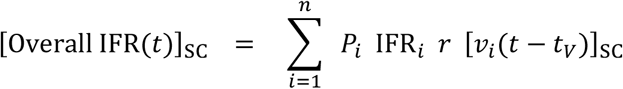

where:

- *t* is the day
- SC refers to the vaccine allocation scenario under consideration, i.e., Actual Allocation (AA) or Equal Allocation by Age (EAA) or Equal Allocation by Population (EAP).
- The summation is across population cohorts *i*based on 5-year-wide age groupings (i.e., 0-4 years, 5-9 years, etc. up to 100+ years old).
- *P*_*i*_is the population of cohort *i*(as a % of the total population) – per estimates, for each country in the world, of population by age group for 2021 made by the United Nations *[10]*.
- IFR_*i*_is the infection fatality ratio for people in the cohort *i*– which are taken from estimated IFRs for each age group from O’Driscoll et al. *[11]* and assumed to be the same for each country.
- *r*is the factor by which mortality is changed due to vaccination – assumed to be 0.1, i.e., a 90% reduction in risk of death, for all age cohorts *[12]*.
- [*v*_*i*_]_SC_ is the percentage of the population in cohort *i* that is vaccinated (under the vaccine allocation scenario SC).
- *t*_*V*_ is the average number of days between receiving a vaccine and gaining immunity due to vaccination.

The values of [*v*_*i*_]_SC_ for each population age cohort are calculated in two steps. First, determine the total percentage of the population that is vaccinated on the particular day, which is determined by the scenarios for the global allocation of vaccines across countries:

- *Actual Allocation (AA):* In this scenario, the actual number of vaccine doses divided by 2 is used as an estimate of the total number of people vaccinated.
- *Equal Allocation by Age (EA):* In this scenario, the age of the youngest person vaccinated is calculated from the total number of vaccines doses worldwide divided by 2 and the global population in each age cohort, and the number of people vaccinated in any country is the number of people above this age.
- *Equal Allocation by Population (EP):* In this scenario, the number of people vaccinated in any country is equal to the country’s population multiplied by the proportion of the global population that was vaccinated on that day (determined by the total number of vaccines doses worldwide divided by 2).

Second, calculate the percentage of each age cohort in the country that is vaccinated. Each country has organized its vaccination rollout differently *[3]*, but we use simplified sequences of vaccinations are for the purposes of these model calculations – and test for how sensitive the results to alternative assumptions. Two types of rollout schedules are used: (1) vaccinations are sequenced by 5-year-wide age cohorts, starting with the oldest, up to an assumed maximum uptake in each cohort; and (2) vaccinations are provided first to people above a given threshold age, and then to people below the threshold age, up to an assumed maximum uptake for the groups above and below the threshold age. For the results presented in Figures 2 and 3, HICs and UMICs are assumed to have the first schedule, and LMICs and LICs are assumed to have the second schedule; the maximum uptake is assumed to be 80% for all countries and all cohorts; and the threshold age in the second schedule is assumed to be 50 years for all applicable countries. It is assumed that each country would have used the same vaccination sequence even if the total number of doses it obtained from the global allocation of vaccines had been different than the actual number of doses received.

For each country, and each day, we estimate the number of reported deaths in the EAA and EAP scenarios, from the actual number of reported deaths, using the following formula:

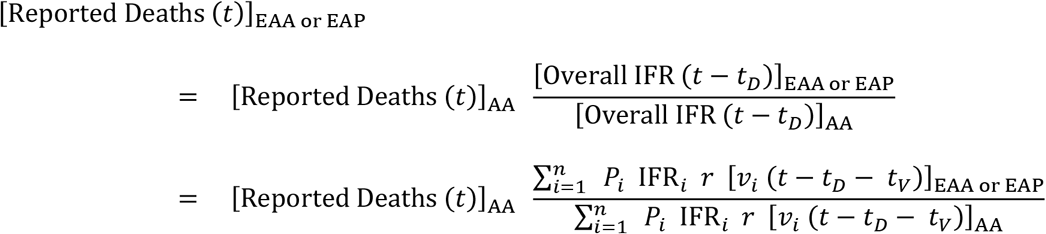

Data for actual reported deaths come from statistics reported by the World Health Organization or in the Our World In Data dataset *[1]*. Deaths on day *t*are caused by infections that occur on average *t*_*D*_ days previously and hence depend on the IFR on day *t*− *t*_*D*_. Vaccination generates immunity to the virus at an average of *t*_*V*_ days after receiving the dose. Hence, deaths on day *t*are affected by the number of people vaccinated on day *t*− *t*_*D*_ − *t*_*V*_, i.e., *t*_*D*_ + *t*_*V*_ days earlier. This gap is assumed to be about 30 days, based on estimates that deaths occur at an average of 20-25 days after infection *[14]* and advice from health authorities that people have full protection at 7 to 14 days after vaccination *[13]*.

In many countries, the real number of deaths from COVID-19 is much more than reported. Hence, the real number of deaths from COVID-19 is related to the reported deaths, for any of the scenarios (SC = AA or EAA or EAP) by a death detection ratio (DDR), i.e.:

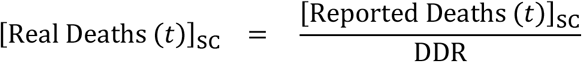

Note that the ratio of the estimates for real numbers of deaths between one scenario and another is independent of the death detection ratio:

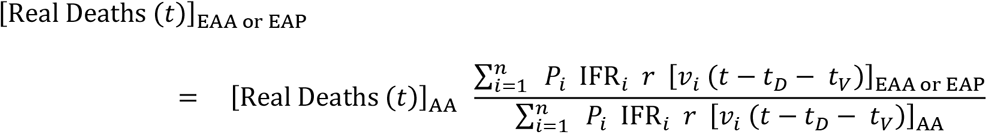

For most countries, we determine the DDR by dividing reported COVID-19 deaths up to 31 October 2021 by the estimates of excess mortality from The Economist on 31 October 2021 *[9]*. For about 100 countries, The Economist drew upon data from the World Mortality Dataset which compares deaths in 2020 and 2021 with expected mortality based on trends prior to the pandemic; for other countries, The Economist used a machine learning model to predict excess mortality based on a range of statistical indicators. For a few countries and territories, the calculations use alternatives to the DDR values derived from The Economist model’s numbers (as accessed on 15 November 2021), as described in *[9]*.

We test the dependence of the findings on the main parameters through a sensitivity analysis. Figure 5 shows the changes in estimated deaths under the two alternative scenarios, when various parameters are changed in turn:

**Figure 5.**
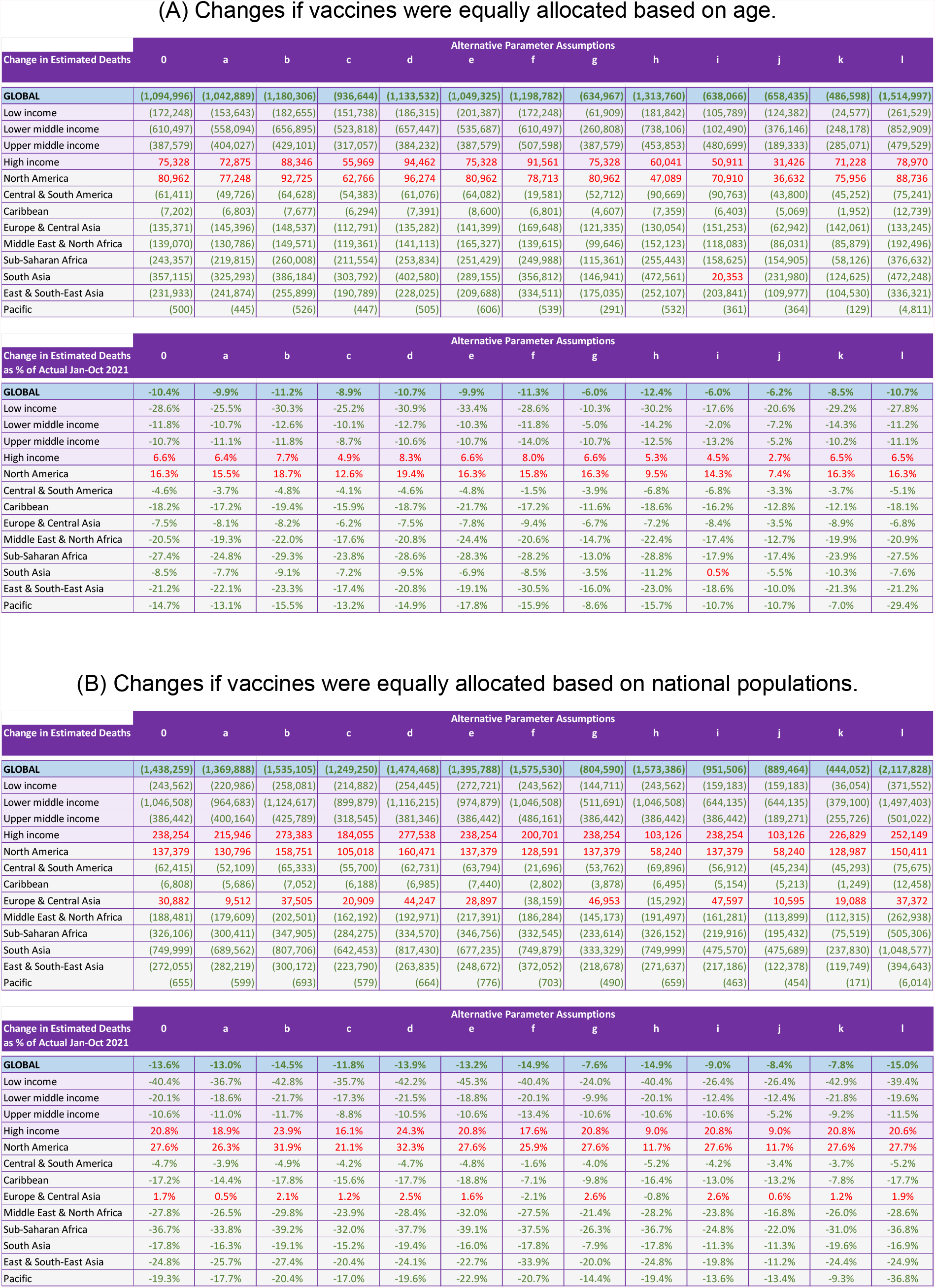
Changes in estimated COVID-19 deaths under the alternative global vaccine allocation rules, for different assumptions regarding key parameters in the calculations. See Figure 4 and the Model Description for explanation of the parameters used for each of the estimates labelled “ 0” and “ a” to “ l”.

a. Using alternative values for the infection fatality ratio at different ages, IFR_*i*_, taken from an earlier analysis by Brazeau et al. *[18]*, which increases slightly less steeply with increasing age.
b. Increasing protection from vaccination to 95%, i.e., decreasing the factor by which mortality is changed due to vaccination, *r*, to 0.05.
c. Increasing protection from vaccination to 80%, i.e., increasing the factor by which mortality is changed due to vaccination, *r*, to 0.20.
d. Changing the average delay between vaccinations and when they determine the reported and actual deaths from COVID-19, i.e., *t*_*D*_ + *t*_*V*_, from 30 to 20 days.
e. Assuming that all countries – including LMICs and LICs as well as HICs and UMICs – have a vaccination schedule in which vaccinations are sequenced by 5-year-wide age cohorts, starting with the oldest.
f. Assuming that all countries – including HICs and UMICs as well as LMICs and LICs – have a vaccination schedule in which vaccinations are provided first to people above a threshold age of 50 years, and then to people below the threshold age, up to an assumed maximum uptake for the groups above and below the threshold age.
g. Assuming that HICs and UMICs have a vaccination schedule in which vaccinations are sequenced by 5-year-wide age cohorts, starting with the oldest, and that LMICs and LICs have a vaccination schedule in which vaccinations are provided to the general population (above 20 years) without sequencing by age.
h. Decreasing the maximum uptake of vaccines from 80% to 50% for HICs only.
i. Decreasing the maximum uptake of vaccines from 80% to 50% for LMICs and LICs only.
j. Decreasing the maximum uptake of vaccines from 80% to 50% for all countries.
k. Using death detection ratios to estimate actual deaths derived from the lower end of the 95% confidence interval for the estimates of excess deaths from The Economist’s model (with the exceptions noted above).
l. Using death detection ratios to estimate actual deaths derived from the upper end of the 95% confidence interval for the estimates of excess deaths from The Economist’s model (with the exceptions noted above).

The major findings do not change in direction – i.e., which global vaccine allocation rule yields the greatest number of lives saved – even if the main parameters vary dramatically from those assumed. However, the magnitude of the differences between vaccine allocation scenarios does change when the parameters are varied. The differences in lives saved are greatest, as a percentage of estimated actual deaths between 1 January and 31 October 2021, when all countries, including HICs and UMICs, are assumed to vaccinate people in just two phases, one for over-50s and the second for under-50s (variation (f), which is realistic for some UMICs but not for most HICs and UMICs), when vaccine uptake in HICs is assumed to be only 50% (variation (h), which is unrealistically low for most HICs), and when vaccination is assumed to reduce mortality by 95% (variation (b), which is likely to be unrealistically high). The differences in lives saved, as a percentage of estimated actual deaths between 1 January and 31 October 2021, are lowest when LMICs and LICs are assumed not to apply any sequencing by age in their vaccination rollouts (variation (g), which does not reflect the stated vaccination policy of any country), when vaccine uptake in LMICs and LICs, or vaccine uptake across all countries, is assumed to be only 50% (variations (i) and (j), which is likely lower than the actual vaccine uptake for these countries in aggregate but could be true for some specific countries due to a combination of difficulties in distributing vaccines and vaccine hesitancy). The differences in lives saved are greatest/smallest, in absolute number, when estimated actual deaths from COVID-19, use the upper/lower end of the confidence intervals from The Economist’s model of excess mortality.

## Supporting information

Data File for COVID-19 Vaccine Allocation Scenarios

## Data Availability

All data used in this study can be freely downloaded from the cited sources. The model outputs, including all the data presented in this article, are contained in the spreadsheet file which accompanies the article.

## Funding

No funding was received to support this work.

## Declaration of interests

The author works at Dalberg Advisors, a consultancy whose clients include multilateral agencies, foundations, international development agencies, governments, companies and NGOs. However, this work was conducted in a personal capacity and was not funded by Dalberg or any of its clients.

## Notes

### Summary of Updates

Added contact email address. Amended the section headings to highlight the main points of each section. No other changes have been made to the text of the article, aside from the section headings.

